# A Rapid Assessment of Mortality Surveillance in Uganda

**DOI:** 10.1101/2024.08.28.24312727

**Authors:** Marie Gorreti Zalwango, Caroline Kyozira, Mariam Nambuya, Martin Bulamu, Allan Muruta

## Abstract

**Background:** Mortality surveillance aids in identifying and addressing their causes allowing health systems to adapt and respond effectively. This rapid assessment aimed to create awareness on the state of mortality surveillance in Uganda, highlight existing gaps and provide recommendations required for an improved mortality system hence the eventual improvement of public health in the country.

**Methods:** An assessment of mortality surveillance in Uganda was conducted from November 2023 to June 2024 through data reviews and plenary discussions engaging various stakeholders in Uganda. Eight (8) workshops/meetings were conducted over a period of eight months to generate information on mortality data sources, processes of data generation and challenges affecting the system. Reports generated from the meetings and workshops were summarized and presented as descriptive narratives. Data from DHIS2 was analyzed using excel and presented using charts and tables.

**Results:** The rapid assessment of mortality surveillance in Uganda highlighted opportunities for improved mortality surveillance through the existence of various sources of data. It was highlighted that 66.9% of the death occur in communities, however, there is a major data completeness gaps where suboptimal data from the community is feed into the national health statistics database (DHIS2) to enable stakeholder analysis and utilization. Furthermore, a number of data quality issues were identified in the health facility generated data where 33% of the deaths occur. These include: data completeness where the national referral specialized health institutes do not feed their data into the national data base, late reporting and the lack of coordination and standardisation of reporting among the various partners.

**Conclusion:** The existence of structures to conduct mortality surveillance in Uganda presents an opportunity for improved mortality surveillance despite the highlighted gaps and challenges. Adoption of strategies aimed to enable the successful implementation of an efficient mortality surveillance program like: strengthening governance and operations of death reporting activities, establishing a clear definition of institutional roles and responsibilities, raising awareness and advocacy at all levels, building technical capacities, improving allocation of resources, and leveraging on shared interests by both implementing and development partners could improve mortality surveillance and the health of the population through utilisation of the generated data.

## INTRODUCTION

Global burden of disease estimates that approximately 131 million (ranging between 126 and 137 million) people died worldwide from all causes over the combined years of 2020 and 2021 [1, 2]. A greater burden of mortality is experienced in Sub-Saharan African countries due to a dual burden of communicable and non-communicable diseases accounting for 36% of deaths in 2019 [3]. In Uganda, the national population as per the 2024 census is 45,935,046 and the life expectancy at birth improved to 67.7 years in 2022 from 65.8 years in 2016[4, 5]. Similarly, the under 5 mortality rate has reduced from 64 deaths / 1,000 live births in 2016 to 52 deaths /1000 live births in 2022[5]. Nonetheless, Uganda has not yet met the Sustainable development goals (SDG3) targets of 12 per 1,000 live births and under-5 mortality to at least as low as 25 per 1,000 live birth; and yet the health and well-being of a country is measured largely by mortality indicators[6].

Population-based data on mortality disaggregated by age, sex, cause-of-death location and multiple other dimensions of inequality are the statistical foundation for public health, however, such information is lacking in almost all developing countries including Uganda, due to inefficient civil registration systems [7]. A multi-country study showed that from 2010 to 2016 African countries scored an average of 8.3% for mortality data accuracy and completeness, as compared to a global average of 46.9% for the same period. Thirty-eight (38;69%) of the 55 countries on the African continent received a zero score [8]. In Uganda, the crude death rate for the year 2024 was approximately 5.9 per 1,000 population yet only 17% of deaths are registered annually [4, 9]. A comprehensive and timely reporting on population health by underlying causes of disability and premature death is crucial to understanding and responding to complex patterns of disease and injury burden over time across age groups, sex, and locations [10].

Monitoring deaths aids in identifying and addressing their causes allowing health systems to adapt and respond effectively. It triggers responses across multiple sectors based on the cause of death, such as: Ministry of Works and Transport for road traffic accidents; Ministry of Agriculture, Animal Industry and Fisheries (MAAIF) for the rising prevalence of food related events among others. Additionally, understanding the reasons why people die, can help comprehend the ways people live to improve health services and reduce premature mortality from effective response to changing epidemiological circumstances[11]. Therefore, the availability, accessibility and utilization of mortality data is fundamentally important for Uganda’s national population health assessment and health development agenda; more so to timely detect and adequately respond to the current and emerging public health threats. This rapid assessment aims to create awareness on the state of mortality surveillance in Uganda, highlight existing gaps and provide recommendations required for an improved mortality system hence the eventual improvement of public health in the country.

## METHODS

### Study setting

Uganda is located in the Eastern part of Africa with a population of 45,935,046. The country’s health system is structured with both the public and private sectors. The public sector has facilities at various levels in a hierarchical form with VHTs and CHEWs at the community level and National referral hospitals as the highest level of care. At these various levels surveillance for mortality is done through documentation and reporting. Mortality surveillance utilizes the existing surveillance human resource as per the health system structure with all human resource mandated to support mortality surveillance to ensure quality in identification, notification, reporting, certification and data use (Figure 1).

**Figure 1.**
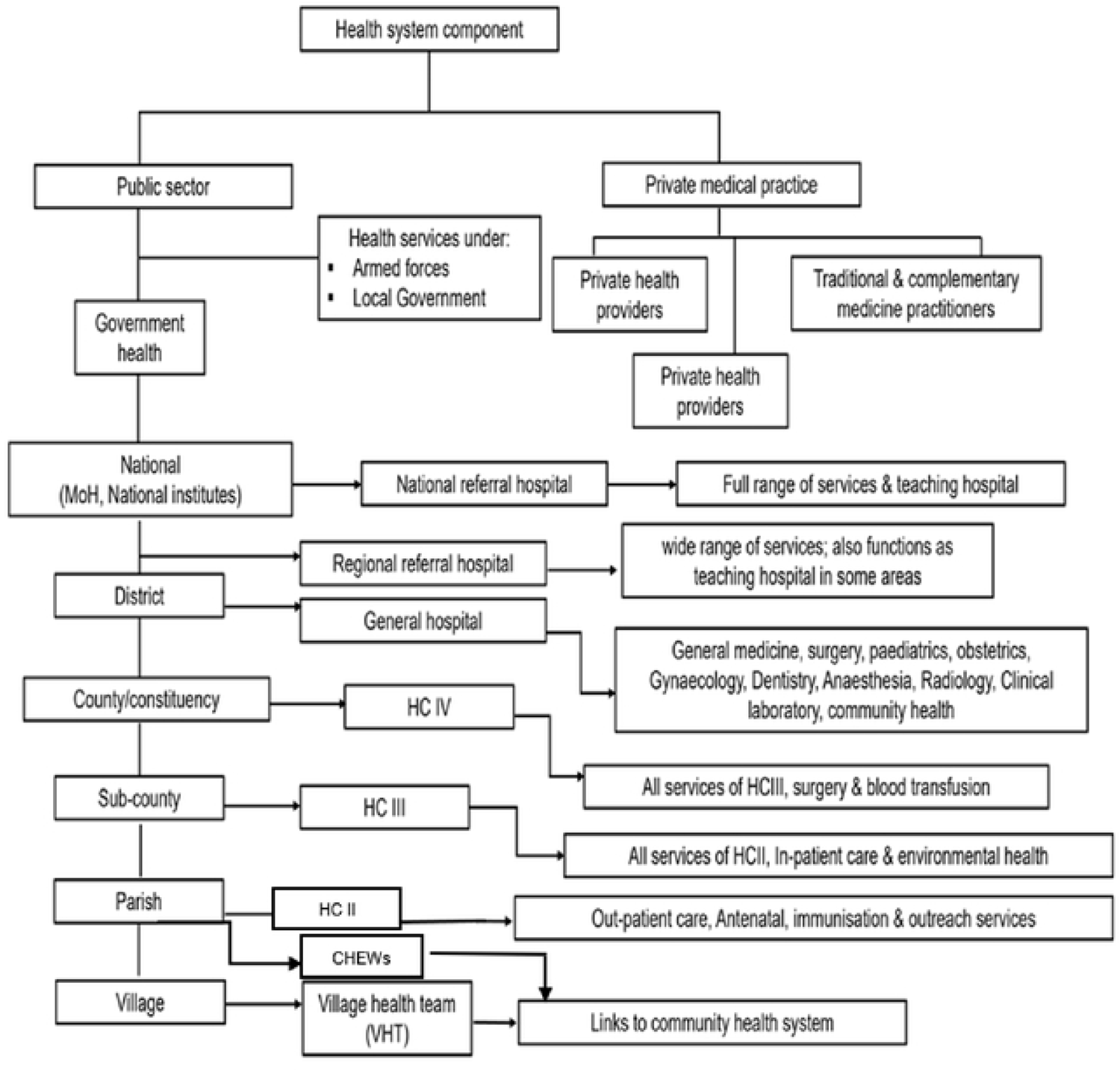
A detailed structure of the healthcare system in Uganda from community to national level

### Study design and data collection

An assessment of mortality surveillance in Uganda was conducted from November 2023 to June 2024 through data reviews and plenary discussions engaging various stakeholders in Uganda. Eight (8) workshops / meetings were conducted over a period of eight months to generate information on mortality data sources, processes of data generation and challenges affecting the system. The stakeholders engaged included but not limited to: key divisions at the ministry of health (the division of health information, Reproductive health division, department of integrated epidemiology, surveillance & public health emergencies, Community Health Department), the National Identification & Registration Authority (NIRA), Semi-autonomous health institutes (Mulago hospital, Uganda heart institute and Uganda Cancer institute), Security agents (Uganda Police-Department of public health, UPDF and Uganda Prisons), Health facility staff at regional and district level, Community health workers, development and implementing partners (CDC, CDCF, WHO, USAID, IDI, AFENET, World Vision and Baylor Uganda), Iganga Mayuge Health Demographic Surveillance Site (IM-HDSS), National Insititute of Public Health, among others.

Through these engagements we explored mortality data sources in Uganda and processes for mortality data flow from the sources to the national database for utilization and finally verified with the mortality data in the district health information system (DHIS2) for accuracy between various reports. The DHIS2 is a web-based reporting system that aggregates data from the health facilities and community; every health facility within the country is responsible for the entry of health data including mortality data within the DHIS2. This system aggregates data by health facility, district, regional and national level and is accessed by various stakeholders to facilitate data use for informed action. The DHIS2 system also houses a national mortality dashboard which was developed to facilitate mortality data visualization and utilization.

### Data Analysis

Reports on mortality data sources and flow processes provided by the various stakeholders were summarized and presented as descriptive narratives and tables. Mortality data in the national dashboard (the various DHIS2 reports) were analyzed using excel and presented as charts and tables.

### Ethical Consideration

We obtained administrative clearance from the Ministry of Health department of Integrated Epidemiology, Surveillance & Public health emergencies. We further obtained verbal consent from all the individuals that participated in the discussions to generate the current status and existing gaps in the country’s mortality surveillance system. Additionally, we obtained routinely collected data from the national health information system which is publicly available for analysis and use to inform public health interventions. This data was aggregated with no individual identifiers. The review was considered non-research as it aimed to highlight and address gaps in the public health sector.

## RESULTS

### General Sources of Mortality Data in Uganda

The implementation of Mortality Surveillance (MS) in Uganda leverages on existing surveillance structures at both national and subnational levels to ensure efficient and sustainable data collection, analysis and use. This section highlights the various sources of mortality data from health facilities, community and various programs supporting mortality surveillance.

### Health facility mortality data

Mortality data is collected from the health facilities using the HMIS registers and reports at both outpatient and in-patient departments. Every death is expected to be reported in the various outpatient and inpatient reports, notified and medically certified. At the outpatient departments, the various registers used to document mortality data include: 1) HMIS 002 - OPD register, 2) HMIS 009 - TB treatment register, 3) HMIS 004-Emergency register and 4) HMIS 006 - Integrated Maternity register. At the inpatient department (IPD), deaths are documented in the respective registers depending on the admitting unit, these units include: 1) HMIS 006: Integrated Maternity register used Maternity, 2) HMIS 011: New-born Inpatient register is used in NICU, 3) HMIS 008: Integrated PNC and Maternity register used in Post Natal ward, 4) HMIS 001L: INR register used in Nutrition units, 5) TB unit treatment register and G expert registers used in TB ward and 6) HMIS 008 Palliative care register used to record palliative care patients.

The HMIS reports used include: the HMIS 033b - Weekly epidemiological surveillance report (Captures deaths for only diseases / events of public health importance) for both inpatient and outpatient departments; the HMIS 105 OPD Monthly Report reports deaths from the various outpatient departments and the HMIS 108 IPD Monthly Report reports deaths from all inpatient departments. Important to note is that some indicators like maternal mortality and neonatal mortality appear in both the monthly outpatient and monthly inpatient reports, a possible cause of duplication.

### Distribution of Mortality Cases per Health Facility Level

The distribution of mortality across different healthcare levels shows a significant variation. Regional and General hospitals report the highest mortality rates with HC IIs reporting the lowest mortality. Important to note is the contribution of private clinics, HC IIs and HC IIIs to mortality (Table 1) yet these facilities are not equipped with medical officers to medically certify these deaths.

**Table 1:**
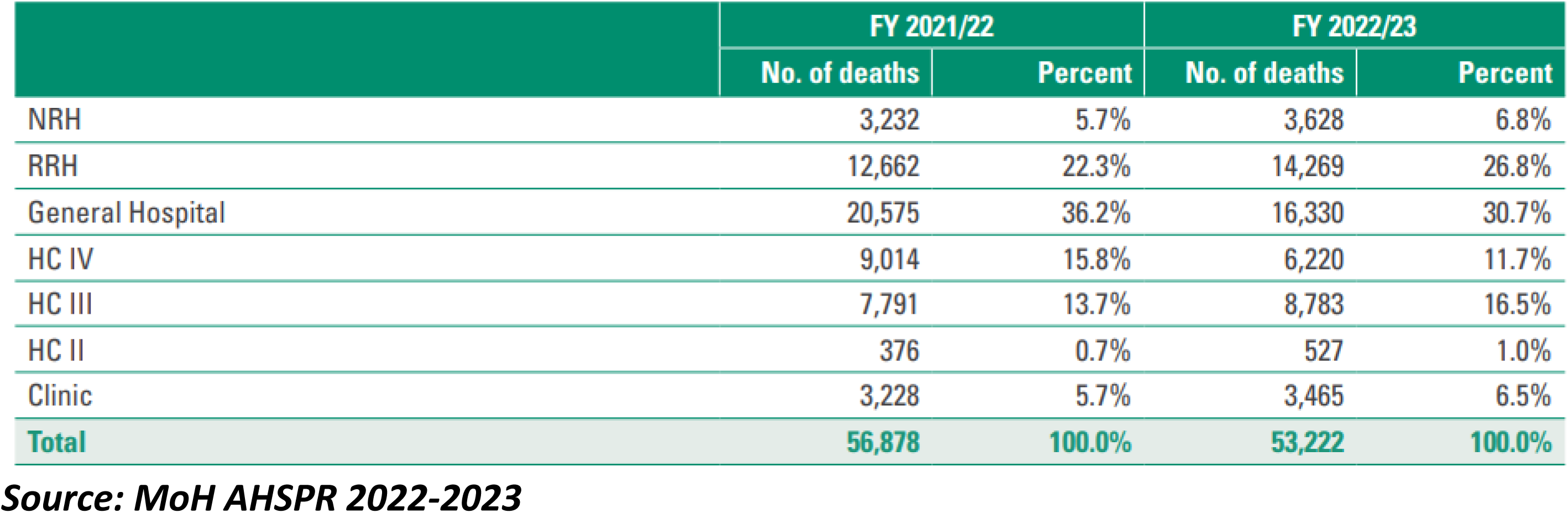
Mortality statistics by health facility level in Uganda, FY 2022-2023.

|  | FY 2021/22 |  | FY 2022/23 |  |
| --- | --- | --- | --- | --- |
|  | No. of deaths | Percent | No. of deaths | Percent |
| NRH | 3,232 | 5.7% | 3,628 | 6.8% |
| RRH | 12,662 | 22.3% | 14,269 | 26.8% |
| General Hospital | 20,575 | 36.2% | 16,330 | 30.7% |
| HC IV | 9,014 | 15.8% | 6,220 | 11.7% |
| HC III | 7,791 | 13.7% | 8,783 | 16.5% |
| HC II | 376 | 0.7% | 527 | 1.0% |
| Clinic | 3,228 | 5.7% | 3,465 | 6.5% |
| **Total** | **56,878** | **100.0%** | **53,222** | **100.0%** |

**Source: MoH AHSPR 2022-2023**

### Notification and Medical Certificate of Cause of Death (MCCOD)

In Uganda, the HMIS Form 100 is for notifying all deaths that take place within a health facility as well as medical certification of cause of death. In the event of death, a medical officer completes the HMIS form 100 according to the World Health Organization’s (WHO) International Classification of Diseases (ICD) standards after which it is entered into the national HMIS / DHIS 2. In 2017, the Ministry of Health and NIRA undertook several measures to strengthen Mortality Surveillance and Civil Registration and Vital Statistics (CRVS) in Uganda. From 2019 to 2022, MOH and NIRA trained a team of health workers from 14 RRHs, 51 General district hospitals, and 115 HC IVs in the international classification of diseases (ICD 11) and medical certification of cause of death (MCCOD) and also harmonized tools for MCCOD including developing the ICD 11 app in DHIS2. Despite these efforts to build capacity for mortality notification and medical certification, these remain low with notification at 19.6% and medical certification at 13.8% according to the mortality dashboard, June 2024 (Figure 2a & 2b).

**Figure 2a:**
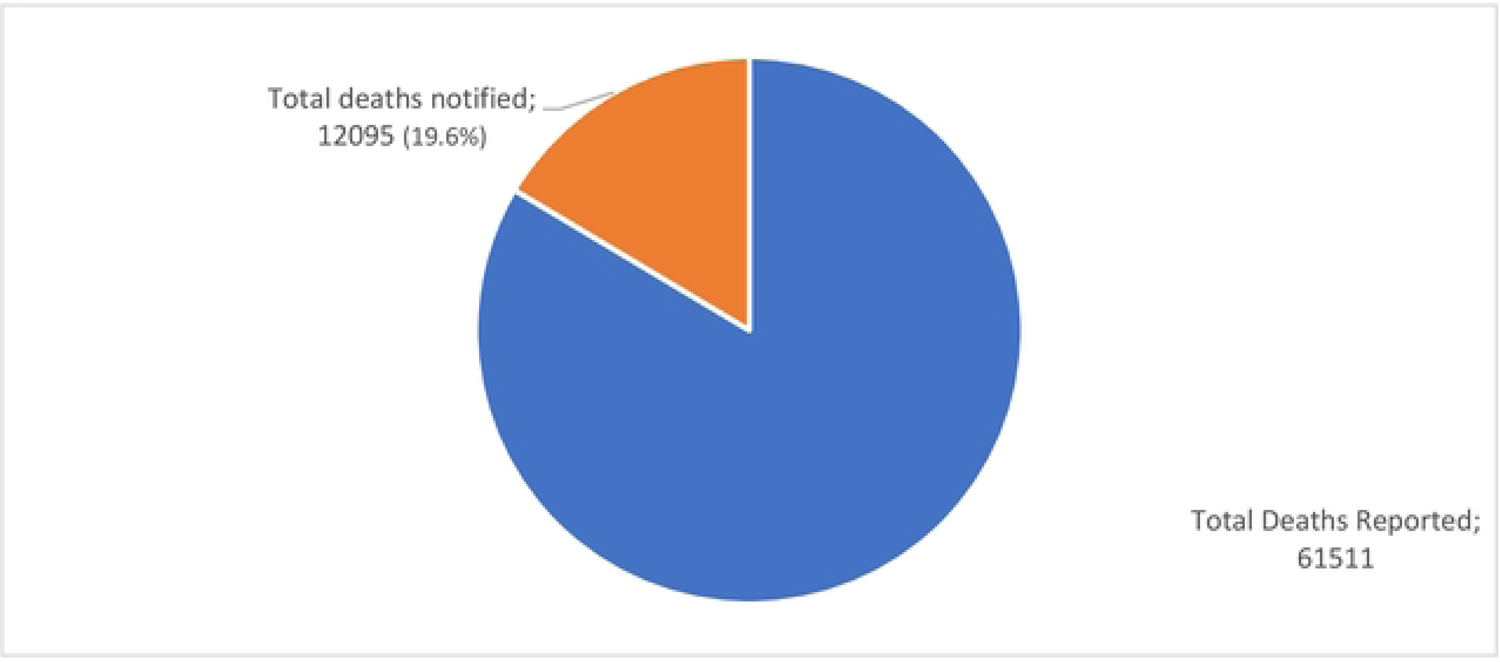
A chart with total mortality cases reported and notified for Uganda, July 2023-June 2024 (*Source: DHIS2*)

**Figure 2b:**
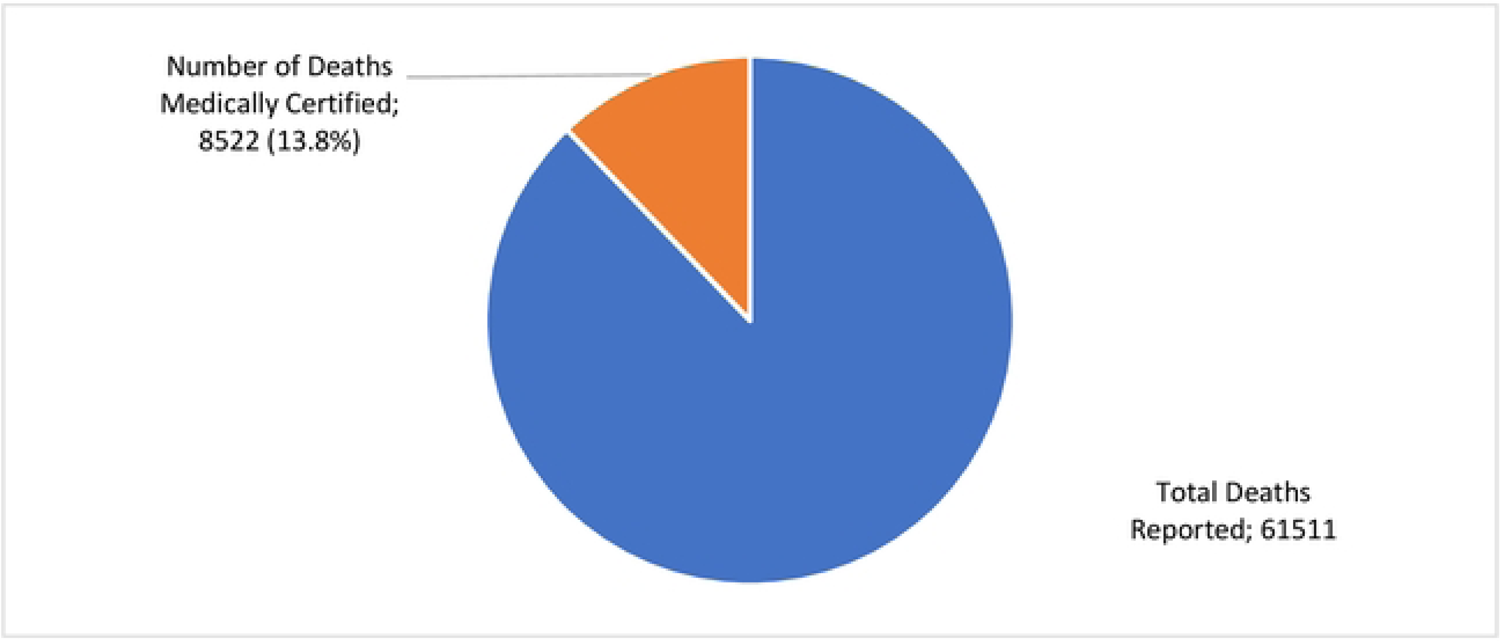
A chart with total mortality cases reported and medically certified for Uganda, July 2023-June 2024 (*Source: DHIS2*)

### Community mortality data

In Uganda, 66.9% of deaths occur within the community [12]. However, mortality data at community level is sub optimally captured under 2 fronts; through the VHTs to health facilities using HMIS 097 and through community local authorities to NIRA using form 12.

### Routine community mortality surveillance

Deaths that occurs in the community is notified by volunteering community health workers/ VHTs and local administrative authorities. VHTs complete the paper-based HMIS 097 report and submit it quarterly to the respective health facilities for entry into the HMIS. However, these reports only capture under 5 and maternal mortalities. Completing reports in the paper-based system is laborious and compounded by challenges such as excessive print volumes, errors in data aggregation, and difficulties in deciphering poor handwriting, undermines the accuracy and timeliness of data and follow-up procedures. Notably, the reporting rates of VHT reports are low at 33% as per the April-June quarter 2024 (Figure 3).

**Figure 3:**
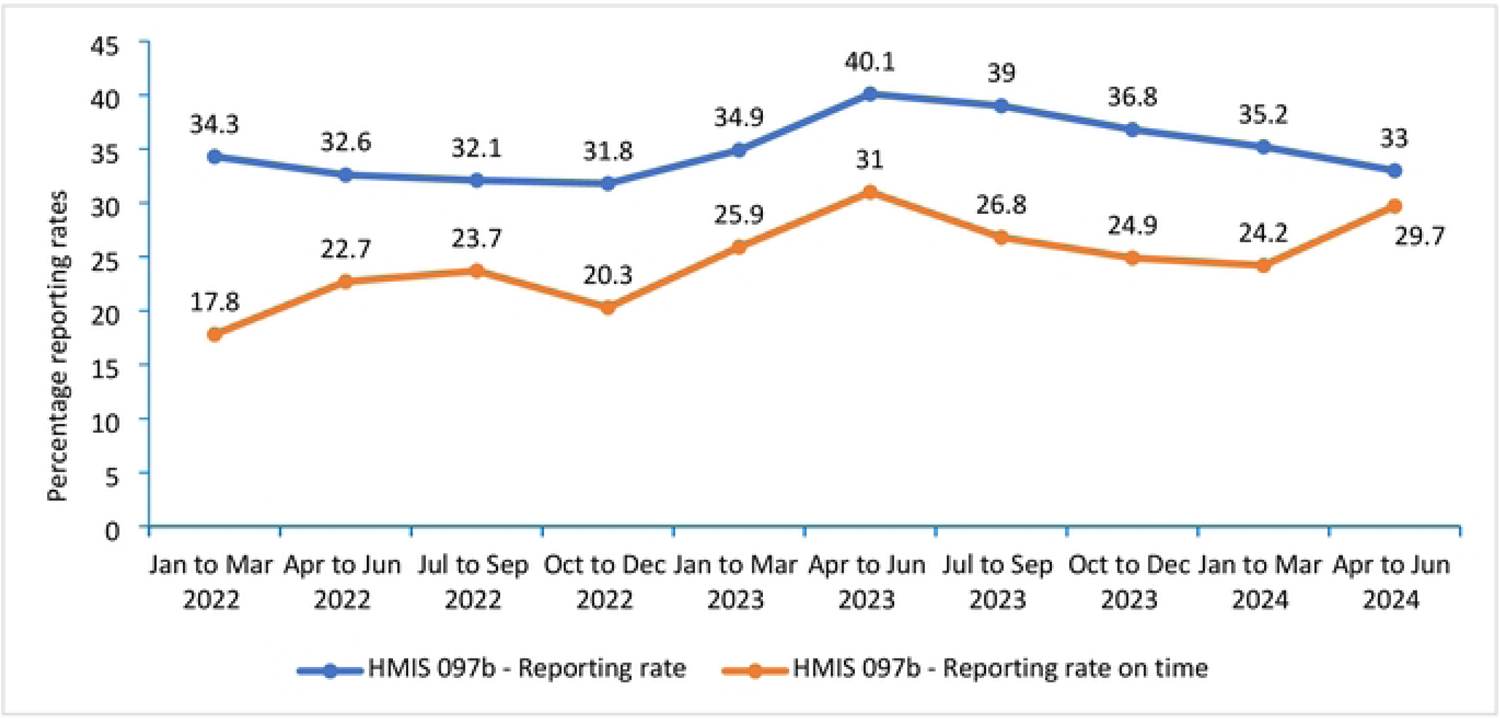
Trends in VHT reporting and timeliness of reports in Uganda, Jan 2022 to June 2024

### Electronic community health Information system (eCHIS)

The eCHIS is a digital platform crafted to assist community health workers in capturing and reporting of health-related information. It covers a spectrum of services including mortality surveillance, Maternal and Child Health (MCH), elements of infectious diseases and care protocols among others.

By April 2024, eCHIS was operational in 17 districts, providing support to more than 11,000 VHTs and 160 Community Health Extension Workers (CHEWs). There are ongoing initiatives to extend its reach nationwide, encompassing both VHTs and CHEWs.

### Mobile Vital Registration System (MVRS) by NIRA

The registration of every death is free and compulsory in Uganda. In March 2014, the country rolled out MVRS by the national Identification & Registration Authority (NIRA). MVRS is currently operational in 135 hospitals and 117 district-level registration centers. *At health facilities, the r*egistration of deaths occurs upon notification using a medical certificate of cause of death (HMIS 100) by a health worker trained to use the MVRS system. Health facilities without MVRS capture death data using the HMIS 100 and once it is entered into the DHSI2, these details are picked on the other end by MVRS. At community level, once the community leaders receive information about the death of an individual, they proceed to notify NIRA using form 12. This form is available at the respective NIRA sub-county offices within the district. Upon completion, the variables on the form are entered into the MVRS for registration and certification.

### Verbal Autopsy

By 2024, VA in Uganda has been implemented at a small scale; a case in point is the Iganga-Mayuge HDSS where probable causes of death at community level are identified and communicated to NIRA for issuance of a death certificate. Verbal autopsy (VA) is a valuable method used to determine the cause of death through interviews with the deceased person’s next of kin or caregivers. These interviews involve a standardized questionnaire to gather details on symptoms, medical history, and the circumstances leading to death. The primary goal of verbal autopsy is to describe the causes of death at the community level or population level in areas where there is no medical certification of deaths or it is not yet well-established. Healthcare professionals or algorithms then analyze this information to identify the likely cause of death.

### Other possible sources of community mortality data

Various sources of mortality data exist at community level. These include: places of worship, traditional leaders, security organs, Traditional Birth attendants (TBAs), funeral homes, cemeteries, police, prisons, post mortem reports and city mortuaries that capture deaths by accidents, suicide and deaths by killing among others. However, currently, these do not feed directly into the national reporting system.

### Census and surveys

A national systematic collection of social demographics, and economic data about all individuals within a specific geographical area at a particular point in time is crucial for government planning, resource allocation, and decision-making. This is through censuses which are done every 10 years, and surveys (Uganda Demographic Health Surveys) that are done every after 5 years among others.

### Health Demographic Surveillance Sites like the Iganga-Mayuge Health & Demographic Surveillance Site (IMHDSS)

The Iganga-Mayuge Health and Demographic Surveillance Site (IMHDSS) which is managed by Makerere University Centre for Health and population research (MUCHAP) was set up in 2005. The HDSS sites collect social demographic data on all individuals in the defined area four times a year. In addition to demographic data, the HDSS also conducts verbal autopsies on all deaths to determine the cause of deaths and related factors like health seeking behaviors and access to health services. Data obtained is comprehensive covering both facility and community deaths, however, they cover limited geographical areas.

### Program specific mortality surveillance

A number of programs within the Ministry of Health conduct mortality surveillance as a mechanism for quality improvement within the departments. These include the maternal and child health (MCH) department maternal and perinatal death Surveillance and Response (MPDSR), TB and HIV programs and malaria. However, there is no unified platform of sharing this information across programs.

### Current Mortality Data flow in Uganda

For effective utilization of mortality data by government and stakeholders, mortality data from the various sources should flow to the central depository. In Uganda however, not all the data collected reaches the national data base for effective utilization (Figure 4).

**Figure 4:**
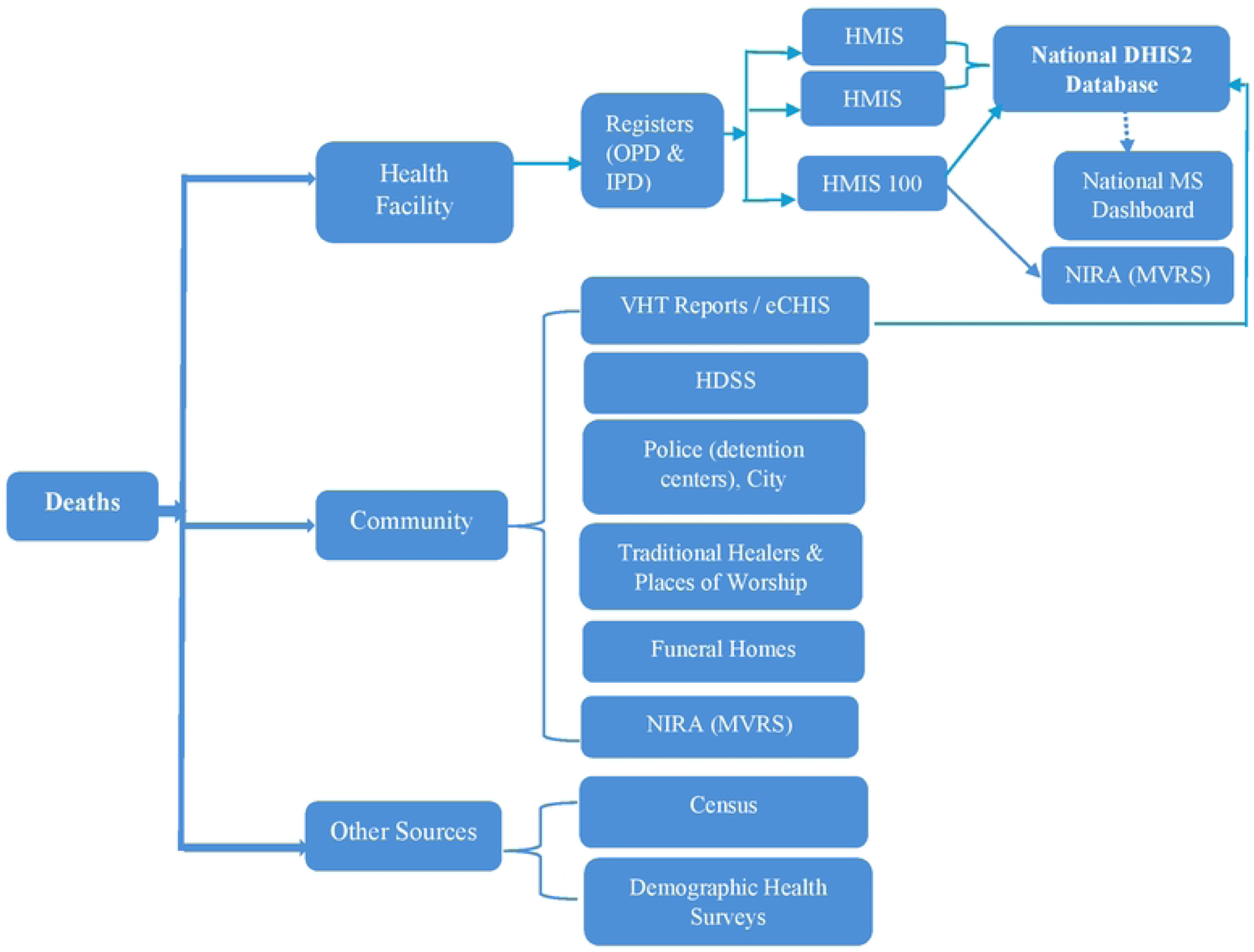
Mortality data flow in Uganda highlighting gaps in mortality reporting from various sources to the national level, 2024

### Data Quality Issues

The quality of data determines the extent at which it can be utilised. To inform decision making, data must be timely, accurate, complete and relevant. Various initiatives have been implemented to improve data quality for MS in Uganda and these include: digitizing CRVS systems, building capacity among health workers and data collectors and engaging communities in death reporting among others. Despite these efforts, significant challenges remain and continued investments and collaboration are needed to ensure informed decision making and improved outcomes.

### Data Completeness

Mortality data utilization at national level is limited by the incompleteness where many deaths remain unreported as well as under reported.

### Mortality data management and reporting by the national health institutes (Uganda Cancer and Heart institutes)

National health institutes are semi-autonomous bodies and among these is the Uganda cancer institute and the Uganda heart institutes. It was noted that not all mortality data from the 2 national institutes is reported into the national HMIS /DHIS2 system. The institutes have parallel institute customised reporting systems that capture data for utilisation by their funders to inform planning. This data is missed out for utilisation by the Ministry of Health as its not captured by the national HMIS system.

### Unreported mortality data

Death data from various facility and community sources (palliative care homes, some private health facilities, places of worship, traditional leaders, security organs, etc…) is often not captured in the national HMIS system. This underreporting undermines the burden of mortality in Uganda hence gaps in planning and strategizing.

### Uncoordinated reporting

Mortality data from a number of mortality surveillance implementers / projects is reported in parallel structures and does not get aggregated by these bodies for a unified national picture.

### Data Timeliness

Timely reporting of mortality data is important for surveillance. Reporting should be made within 7 days to guide timely detection of public health events as per the 7-1-7 matrix. Uganda has a weekly surveillance report aimed at providing timely data on priority events of public health importance; however, timeliness of this weekly surveillance report remains a challenge (Figure 5).

**Figure 5:**
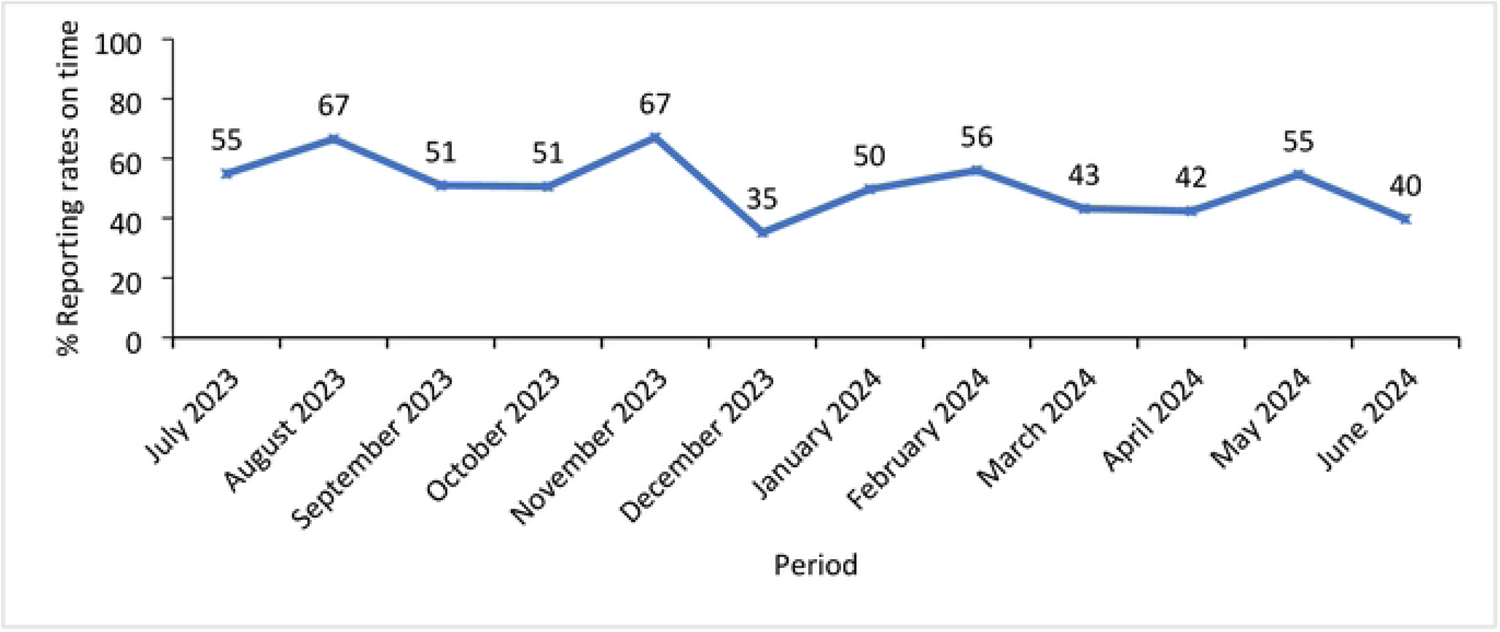
Reporting rates for the HMIS 033b weekly surveillance report in Uganda, July 2023 – June 2024

### Unclear indicator definitions

The monthly outpatient report has an indicator “Death in OPD” and “Total deaths in the emergency unit at OPD”, however, it’s unclear whether death in OPD includes all OPD deaths or it excludes deaths reported from the emergency unit. This lack of clarity in indicator definitions may cause duplication or omission of mortality data affecting its quality.

### Possible duplication in reporting of mortality data

Exploration and review of a few indicators and system data revealed duplication of mortality data in the monthly report. Maternal and neonatal mortality indicators are reported in both outpatient and inpatient monthly reports with the same data source document “integrated maternity register”.

### Mortality data discrepancy between reports

Mortality data at health facilities is reported on weekly through the weekly surveillance report (HMIS 033b report). This report captures outpatient and inpatient deaths within a specified week. Ideally, the monthly mortality data should be comparable to the weekly mortality data for the same indicators (HMIS033b = HMIS105 + HMIS108), however, the assessment of these reports revealed discrepancies. Using maternal mortality as an example, the weekly report (HMIS 033b) reported less deaths compared to the 2 monthly reports. Furthermore, the monthly outpatient report recorded more maternal deaths than the weekly report and monthly inpatient reports. This highlights the need for a deep dive into maternal mortality data reported for harmonization, improved reporting and evidence-based decision making (Figure 6).

**Figure 6:**
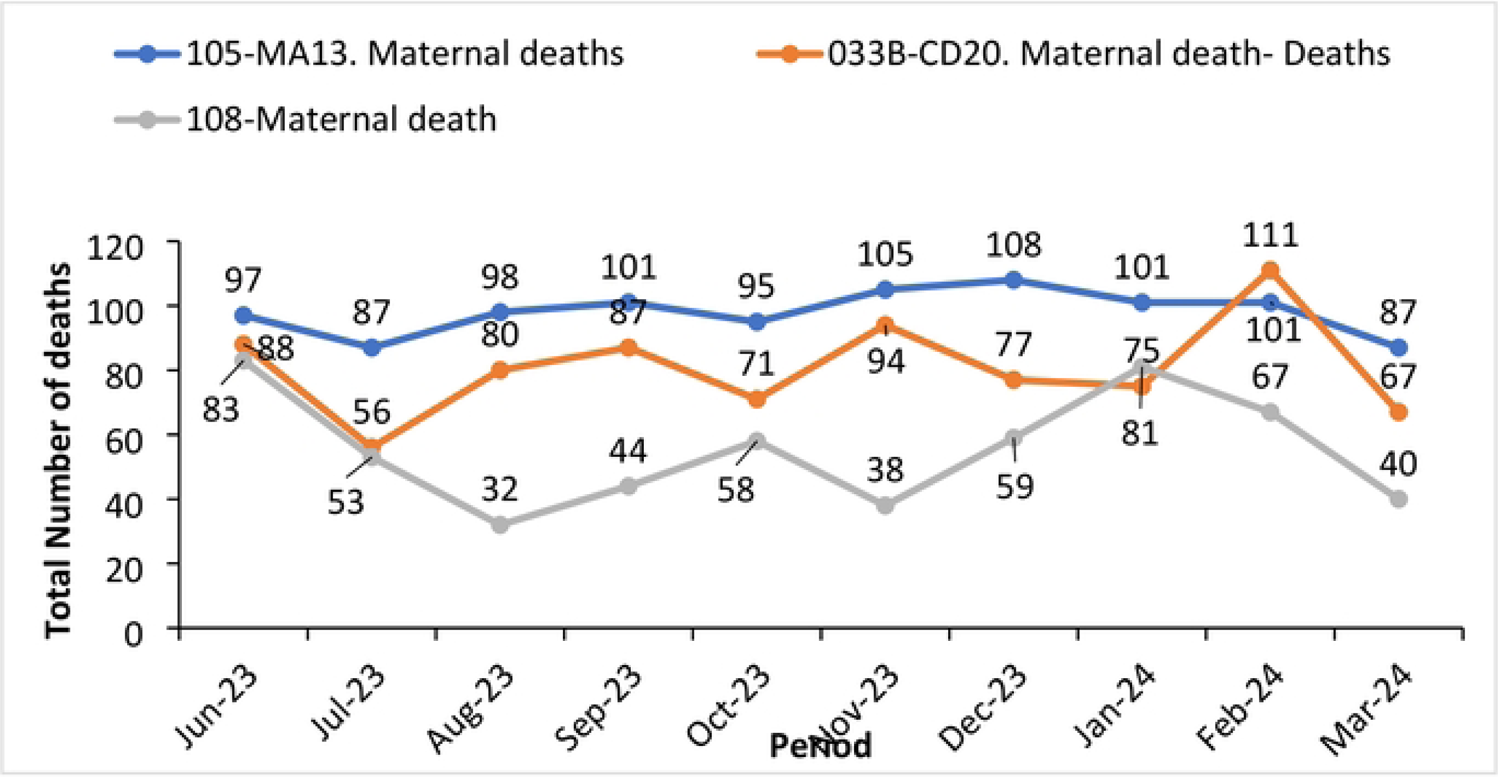
National maternal mortality trends for various in Uganda from June 2023 to March 2024

### Mortality surveillance dashboard

The Ministry of Health designed a mortality dashboard as a one stop center for visualization of mortality data. However, this is for only data coming through DHIS2. During assessment of the data in the MS dashboard, it was noted that there were discrepancies between the reported deaths and the total deaths from the outpatient and inpatient reports in the same system. There is need to verify indicator sources for the mortality dash board to ensure data quality and representation of mortality cases reported from the health facilities through the monthly reports (Figure 7).

**Figure 7:**
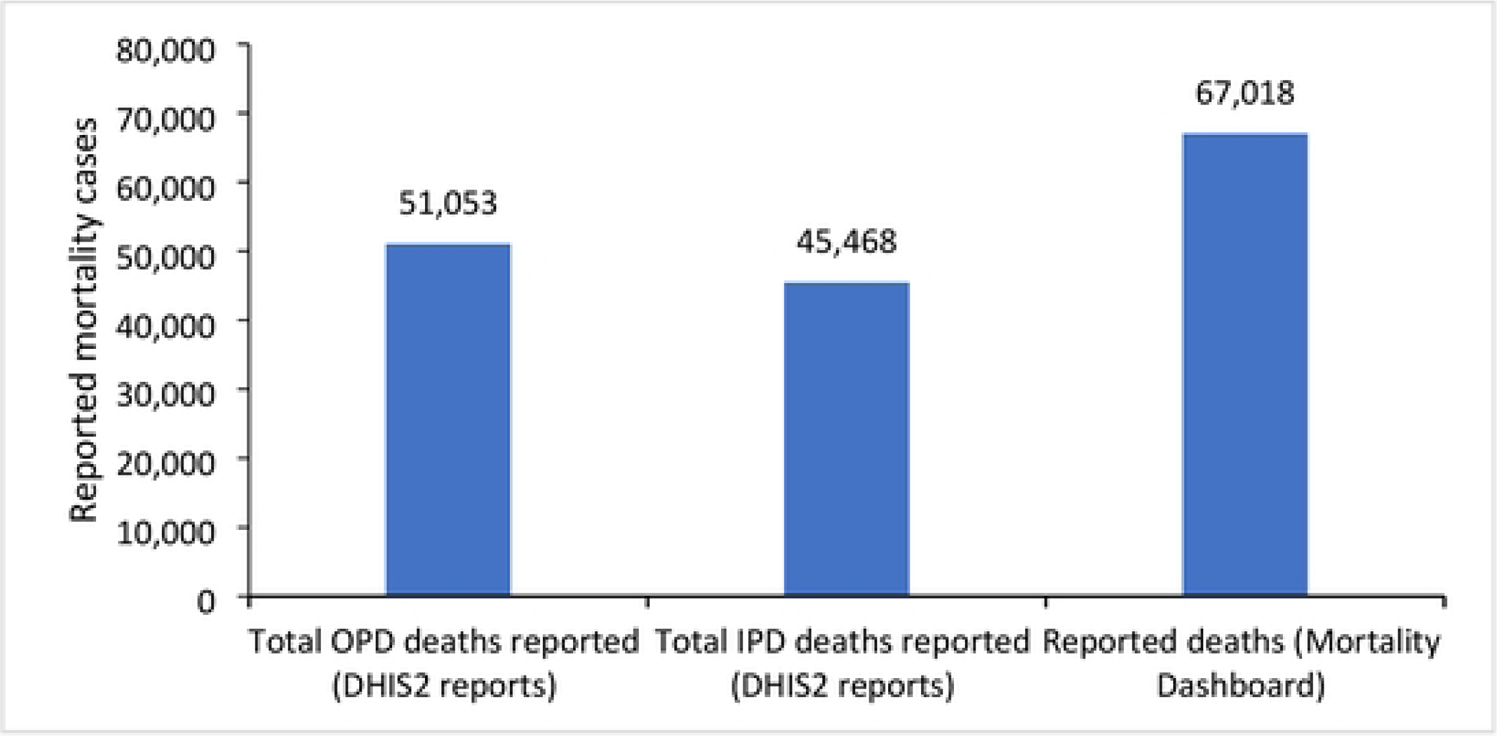
A comparison of mortality cases reported in DHIS2 against mortality cases reported in the national mortality dashboard for the period of May 2023 – April 2024

### Lack of Standardization

The absence of standardized guidelines for mortality reporting can lead to variations in how data are collected and reported, making it difficult to draw comprehensive conclusions or formulate effective health policies.

## DISCUSSION

The rapid assessment of mortality surveillance in Uganda highlighted opportunities for improved mortality surveillance through the existence of various sources of data. The assessment further highlighted a major gap in the identified sources of data feeding into the national health statistics database (DHIS2) to enable stakeholder analysis and utilization. Additionally, the review presented existing data quality issues from the generated data at the national level. These gaps highlight areas in which focus can be drawn to improve data quality including timeliness and completeness. Timeliness is of essence in surveillance as it enables timely decision making.

The report highlights various sources of mortality data, however, there is limited data utilization which is attributed to the incomplete nature of the data. This finding is similar to other countries within Africa where mortality data from various sources has been found to have numerous accuracy and completeness challenges[8]. Countries with fully developed national CRVS systems rely on them for provision of high-quality data. However, in countries like Uganda where a fully viable CRVS system is non-existent, triangulation of a range of data sources can serve as an alternative for compiling mortality data as each of these systems complement each other. The censuses and surveys conducted periodically though not adequate to make timely policy decisions, their data is quite accurate; HDSS provide continuous data on death events and their causes, however, their geographical scope is limited. The HMIS provides continuous fact of death, and cause of death data, however, the data is of aggregate nature and faces challenges with reporting community-based data which is grossly incomplete and yet the country has over 60% of deaths occurring in the community [2]. Furthermore, leading producers of mortality data like specialized health institutions including the Uganda Heart Institute and Uganda Cancer Institute among others do not feed their mortality data in the national database for utilization. Other countries have implemented community-based mortality surveillance through the use of VHTs to collect data, submit it to the sub-national level for verification and later to the national level by the VHT supervisors[13]. Adoption of effective strategies customized to the Ugandan context could improve data capture from the various sources currently not reporting for improved data collection and utilization.

Good public health decision-making is dependent on reliable and timely data on births and deaths, including cause of death. In Uganda, the cause of deaths is determined by a medical officer; however, based on the staffing norms for the country, HC IIs and HC IIIs are not staffed with medical officers yet deaths happen at these levels. Similarly, private clinics are occasionally not staffed with medical officers. In a study conducted in Uganda to understand the application of the workload indicators of staffing need method, HCIV and hospitals which are considered for medical officer staffing had a staffing gap of 39-42%[14]. In another report by IntraHealth International, they highlighted a shortage in staffing for mainly doctors among Ugandan health facilities [15]. This gap is reflected in the low rate of medical certification of deaths in Uganda. Verbal Autopsy has been found helpful in determining the causes of death in populations not served by official medical certification of cause at the time of death in other countries[16]. Adoption of this strategy could address challenges of determining cause of death in health facilities not staffed with medical officers and those occurring in the community.

### Study limitations

Our study relied on self-reports from the various stakeholders engaged in the workshops and dealers for most of the data on mortality surveillance which might have led to social desirability bias. Nevertheless, we think social desirability was minimized through triangulation of data from the national health information system. We also endeavored to emphasize that the results obtained were aimed to improvement of the systems to encourage disclosure of the actual implementation process.

## Conclusion

Mortality surveillance in Uganda is implemented by various stakeholders and numerous sources of data exist. However, for a number of sources, mortality data does not reach the national system for utilization to inform public health action. Furthermore, a number of data quality issues exist like the data completeness where a number of data sources do not completely report to the national system including the high-volume national institutes, lack of standardisation and uncoordinated reporting among the various implementing partners, untimely data submission which affects timely decision making and unclear indicator definitions affecting data quality and utilisation. Despite these gaps, the existence of surveillance structures to conduct mortality surveillance in the country presents an opportunity for improved mortality surveillance and improved health of the population through utilisation of the generated data.

### Recommendations

The recommendations below have been suggested to support the strengthening of the existing structures for improved mortality data collection, analysis and utilization to accurately inform public health actions. These include: Strengthening the governance and leadership structures for mortality surveillance within the country should be prioritized for improved coordination of all stakeholders. To this end therefore, the MS stakeholders in Uganda should jointly design and disseminate mortality surveillance guidelines. They should further institute measures for adherence to the set guidelines to ensure generation of complete, reliable and timely data on all-cause mortality so as to monitor, understand, and address mortality and its underlying causes for public health action.

The MS stakeholders should strengthen notification and registration of deaths at community level through adoption and customization of strategies implemented in other countries for improved accuracy of mortality data in the country for example verbal autopsy. This will support the determination of probable cause of death for community deaths and deaths in low level health facilities without medical doctors.

Ultimately, the country should adopt proposed strategies in the continental framework for strengthening mortality surveillance which are aimed at enabling the successful implementation of a robust mortality surveillance program. These strategies include: strengthening governance and operations of death reporting activities, establishing a clear definition of institutional roles and responsibilities, raising awareness and advocacy at all levels, building technical capacities, improving allocation of resources, and leveraging on shared interests by both implementing and development partners.

## List of abbreviations

MS: Mortality Surveillance

DHIS2: District Health Information System Version 2

HMIS: Health Management Information System

CHEWs: Community Health Extension Workers

NIRA: National Identification and Registration Authority

CRVS: Civil Registration and Vital Statistics

MVRS: Mobile Vital Records System

DQA: Data Quality Assessment

AFENET: African Field Epidemiology Network

CDC: Centers for Disease Control and Prevention

CDCF: Centers for Disease Control and Prevention Foundation

EVD: Ebola Virus Disease

HC II: Health Center Level 2

HC IV: Health Center level 4

RRHs: Regional Referral Hospitals

UCI: Uganda Cancer Institute

UHI: Uganda Heart Institute

HIV: Human Immunodeficiency Virus

HW: Health Worker

ICD11: International Classification of Diseases Version 11

IDSR: Integrated Disease Surveillance and Response

IEC: Information, Education and Communication Materials

IMHDSS: Iganga-Mayuge Health & Demographic Surveillance Site

LC1: Local Council 1

MAAIF: Ministry of Agriculture, Animal Industry and Fisheries

MCCOD: Medical Certification of Cause of Death

MoH: Ministry of Health

MPDSR: Maternal and Perinatal Death Surveillance and Response

TB: Tuberculosis

UBOS: Uganda Bureau of Statistics

UDHS: Uganda Demographic Health Surveys

UNIPH: Uganda National Institute of Public Health

UPDF: Uganda People Defense Force

UPF: Uganda Police Force

USAID: United States Agency for International Development

VHTs: Village Health Teams

VA: Verbal Autopsy

WHO: World Health Organization

## Declarations

### Availability of data and materials

The datasets and reports upon which our findings are based belong to the Ministry of Health. These data sets can be availed upon request from the Ministry of Health through the mortality surveillance focal person Ms. Caroline Kyozira.

### Consent for publication

Not applicable.

### Funding and Disclaimer

This work was done by the Ministry of Health, together with Partners Infectious Disease Institute, Baylor Uganda, and World Health Organization. The staff of the funding bodies including the mortality surveillance stakeholders participated in the workshops and meetings that generated these results and provided technical guidance in writing the manuscript.

### Authors’ Contributions

MGZ, CK and AM conceptualized the idea, analyzed and interpreted the data and drafted the manuscript. MN, and MB reviewed the manuscript. All co-authors read and approved the final manuscript.

## Acknowledgements

We appreciate the Ministry of Health, the staff for the various stakeholders that participated in the workshops and meetings that generated these findings. In a special way we would like to appreciate the Commissioner IES&PHE for his unwavering support, and the partners that funded this piece of work.

